# Effect of behavioural activation for individuals with post-stroke depression: A systematic review and meta-analysis

**DOI:** 10.1101/2023.03.15.23287337

**Authors:** Sandra Walsh, Engida Yisma, Susan Hillier, Marianne Gillam, Richard Gray, Martin Jones

## Abstract

**Objectives:** Post-stroke depression (PSD) affects one third of stroke survivors and substantially impacts recovery, so it is important to identify effective treatments for PSD. Behavioural activation (BA), a relatively simple intervention, has demonstrated efficacy in the treatment of adults with unipolar depression. However, its use and efficacy in the treatment of PSD has not been documented in a systematic review. This systematic review and meta-analysis considers: What effect does BA have on reducing depression symptoms in individuals diagnosed with PSD?

**Method:** MEDLINE, Embase, Emcare, Cochrane Library, and PsycINFO, were searched for randomised controlled trials (RCTs) published in English, on 13 July 2021. To chart the data, we employed a customized extraction sheet. The Cochrane Collaboration risk-of-bias 2 tool was used to determine study quality.

**Results:** Two papers, written by the same lead authors, met the inclusion criteria. The Communication and Low Mood trial and the Behavioural Activation Therapy for Post-Stroke Depression trial were conducted in the United Kingdom and published in 2012 and 2019, respectively. Meta-analysis showed BA was associated with a decrease in depression symptoms in patients with PSD relative to treatment as usual (standard mean difference (SMD) -0.54; 95% confidence interval (CI) -0.90 to -0.18).

**Conclusion:** BA may be more effective when compared to treatment as usual in reducing depression symptoms in individuals with PSD. Although our meta-analysis found positive effects of BA, the evidence is inconclusive due to the small number of studies. High-quality RCTs are needed to confirm the benefits of BA in PSD.

**Strengths and limitations of this study:**

- This review followed PRISMA guidelines with a protocol published in advance of the review being conducted.
- This is the first review to consider whether BA is effective in reducing PSD.
- We focused on depression as a single primary outcome; other outcome measures, such as quality of life, could have been considered.
- Based on the review questions inclusion was limited to experimental designs. Other designs provide additional insights into the application of BA to PSD that is beyond the scope of this study.
- Our review was restricted to studies published in English and did not examine grey literature; therefore, some studies may not have been included.

## Background

### Impact of post-stroke depression

Approximately 1 in 3 stroke survivors develop depression in the first five years post-stroke^1,2^. Depression negatively impacts stroke recovery contributing to higher mortality or hospitalisation, and reduced quality of life^3^. People with post-stroke depression (PSD) are less likely to engage with stroke rehabilitation programs^4^. The health and wellbeing of informal care givers is also affected by living with someone with PSD^5^.

### Care and treatment of PSD

Many stroke survivors who develop PSD are prescribed antidepressants, however when compared to people with depression not related to stroke, there appears to be poorer efficacy, with some experiencing no improvement in mood^2^. In addition, antidepressants have side-effects, which may have a negative impact on stroke survivors, including an increased risk of having another stroke^6^. It has been reported that generally, most people prefer psychological treatment over pharmacological treatments^7^. Psychological treatments such as Cognitive Behavioral Therapy (CBT) may improve mood in PSD. Wang et al.^8^ in a meta-analysis of 23 randomised controlled trials (RCTs), involving 1972 participants, identified that CBT for PSD was associated with positive effect on symptoms of depression. However, CBT can be problematic to deliver. Typically, professionals require extensive training to practice CBT and a high level of clinical skill is required to deliver CBT proficiently^9^.

### Behavioral Activation as a candidate treatment for PSD

Behavioral Activation (BA), a component of CBT, is easy to deliver and could be a candidate psychological intervention for individuals with PSD. It supports the person to engage in meaningful activities and teaches skills to notice changes in mood and its relationship with these activities. Mastery of these activities provides fulfillment and reward. The aim of therapy is to help the patient schedule activity which is inherently rewarding. This engagement with rewarding activity may be particularly important for people with stroke who experience diminished physical capability. Training in BA typically takes about five days^9^, and can be delivered by non-specialist mental health professionals ^10^. Typically, BA is delivered face to face over 6-10 ten sessions^9-12^.

BA has been shown to be effective in treating people with depression and may be as effective as CBT. Richards et al.^10^, reported an equivalence trial that compared CBT with a similar dose of BA in 221 people with moderate to severe depression. They reported that CBT and BA were equally as effective. Furthermore, a recent systematic review by Uphoff et al.^13^ examined BA compared with other psychological therapies, medication, or treatment as usual (TAU)/waiting list/placebo for depression in adults. The review included 53 studies with 5495 participants. The authors concluded that BA “may be more effective than humanistic therapy, medication, and treatment as usual, and that it may be no less effective than CBT, psychodynamic therapy, or being placed on a waiting list” (p.2). They also suggested that BA was just as acceptable to the patients considering other comparators, such as CBT and TAU. While the authors suggested concern regarding the confidence of the findings and longer term follow up of participants was warranted, the results suggest that BA is a worthwhile area of investigation for adults with depression.

BA could be suitable for PSD as its aim is to introduce behaviour that promotes mastery, pleasure, and routine tailored to the individual^11,14,15^. For many reasons, including apprehension, fear, or avoidance, post stroke survivors may disengage from activities that were once pleasurable. For example, a person may stop cooking poststroke, an activity they previously found pleasurable, due to concerns of personal safety. Engaging in incremental cooking activities may provide a sense of pleasure and mastery. The focus of BA could also be in engaging in behaviours that promote routine^14,15^. For someone poststroke, this could be important as it may provide the person with a sense of control and regaining skills. Furthermore BA, as a relatively simple intervention^10^, may be well suited to people with cognitive impairment and barriers to communication as a result of stroke.

Currently, there is limited evidence to confirm or refute whether BA would be beneficial for people with PSD. Therefore, there is merit in conducting research to understand the effectiveness of BA as a psychological treatment for PSD.

### Purpose of the review

We conducted this systematic review and meta-analysis to answer the following key research question: What effect does behavioural activation have on reducing depression symptoms in individuals with post-stroke depression?

## Methods

A protocol was developed for this systematic review and published on the Open Science Framework (Weblink: https://osf.io/7kqu3, Registration 10.17605/OSF.IO/7KQU3). This systematic review was conducted in accordance with the Preferred Reporting Items for Systematic reviews and Meta-Analyses (PRISMA) guidelines^16^.

### Inclusion criteria

Our systematic review included RCTs, non-randomized controlled studies, and controlled before and after studies for adult (>18 years) patients with a primary diagnosis of post-stroke depression and who were treated with BA. We included studies published in English and placed no restriction based on sample size, follow-up period, and publication year.

### Participants/population

Adult patients (>18 years of age) who developed depression following stroke.

### Intervention

Behavioural activation, which involves key elements of self-monitoring and activity scheduling. We defined BA as a “behaviourally oriented, time limited psychotherapeutic intervention, including key elements of self-monitoring and activity scheduling.”^17^

### Comparator

Treatment as usual, comparative depression treatment, placebo, or wait list.

### Outcome

The main outcome measure was change in depression as assessed by any standardized measure of depression, such as the Hospital Anxiety and Depression Scale (HADS)^18^ or the Patient Health Questionnaire-9 (PHQ-9)^19^.

### Search strategy

We searched the following electronic databases: MEDLINE, Embase, Emcare, Cochrane Library, and PsycINFO (the search strategy is available from Open Science Framework https://osf.io/7kqu3, Registration 10.17605/OSF.IO/7KQU3).

The population search terms related to stroke included: stroke, poststroke, cerebrovascular, “cerebrovascular accident”. The population search terms related to depression and included: - depression, depressive, “emotional depression”, “depressive symptom”, mood, low mood, depressed, dysthymia, “vascular depression”, “poststroke depression” and “depressive disorder”.

The intervention search terms (keywords) related to behavioral activation included: “behavior* activation”, “behavior* therapy”, “activity schedul*”, “positive reinforce*”, “event schedul*”, “mood monitoring”, “behavio* treatment”, “behavio* intervention”, “behavio* modif*”, and “behavio* psychotherap*”.

No date limits were applied to the database searches and the date of the search was used as the end date limiter (13 July 2021). The references of included studies were examined to identify any further studies that could be included.

### Study selection

We exported all the publications identified in the searches from EndNote^20^ to Covidence^21^. To identify studies that possibly met the inclusion criteria, two reviewers (MJ/ED) independently screened the ‘titles and/or abstracts’ of each study retrieved. Then, the full text of the identified studies was independently assessed for eligibility by MJ/SW. Any discrepancies were resolved by involving a third reviewer (RG).

### Data extraction

We used an extraction sheet to extract data from the included studies. The selected information from each study included (1) basic information such as authors’ name, year of publication, the number of cases in each group, the proportion of men and women, average age, stroke details (side, latency, type and severity), duration of follow-up time, study design including outcome measures (type and timepoints) and full intervention details (type, frequency etc); and (2) statistical data for the primary outcome (post-stroke depression). Two reviewers (MJ/SW) extracted data independently and discrepancies that arose between the two reviewers were resolved by consensus.

We contacted the corresponding author of two papers to clarify whether the intervention was purely BA or whether other components were included, and to establish whether there was an intention to conduct a larger RCT after the initial feasibility trial. There was no response from the author.

### Assessment of risk of bias in included studies

All studies meeting inclusion criteria were assessed for risk of bias using the Cochrane Risk of Bias Tool,^22^ which considers the following domains: (1) risk of bias arising from the randomisation process, including allocation and randomisation (2) risk of bias due to deviations from the intended interventions, including blinding of participants and people delivering the interventions (3) risk of bias due to missing outcome data, (4) risk of bias in measurement of the outcome, including blinding of outcome assessors, and (5) risk of bias in the selection of reported results.

Two review authors (EY, and MJ) independently assessed the risk of bias in included trials and discussed any disagreements with a third review author (RG). All ‘Risk of bias’ data were presented graphically using *robvis* (visualization tool),^23^ and narratively in the text.

### Data synthesis and statistical analysis

We provided a narrative synthesis of the findings from the included studies. A meta-analysis was also conducted to provide a consolidated estimate of the effect of behavioural activation versus treatment as usual on reducing depression symptoms in individuals with post-stroke depression. We synthesized the information regarding reduction in post-stroke depression symptoms following behavioral activation therapy versus treatment as usual by undertaking random effects meta-analysis using inverse variance weighting. We showed the meta-analyzed effect size (standardized mean difference (SMD)) and its 95 % confidence interval (CI)) as an overall estimate. We used the I^2^ to measure statistical heterogeneity. All analyses were conducted using STATA/SE version 17.0 (Stata Corporation, College Station, TX).

### Patient and public involvement

Patients and public were not involved in this study as it was to establish what had been done in the field. The next phase will involve patients in the research.

## Results

As is show in Figure 1, 765 references were initially identified and 275 duplicates removed, leaving 490 studies to screen at the title and abstract level. After this initial screening, 479 studies were excluded, and 11 studies were assessed for full-text eligibility. Nine studies were excluded as they used the wrong intervention (7), wrong outcomes (1) or were a duplicate (1). Two studies^24,25^ were included for data extraction.

**Figure 1.**
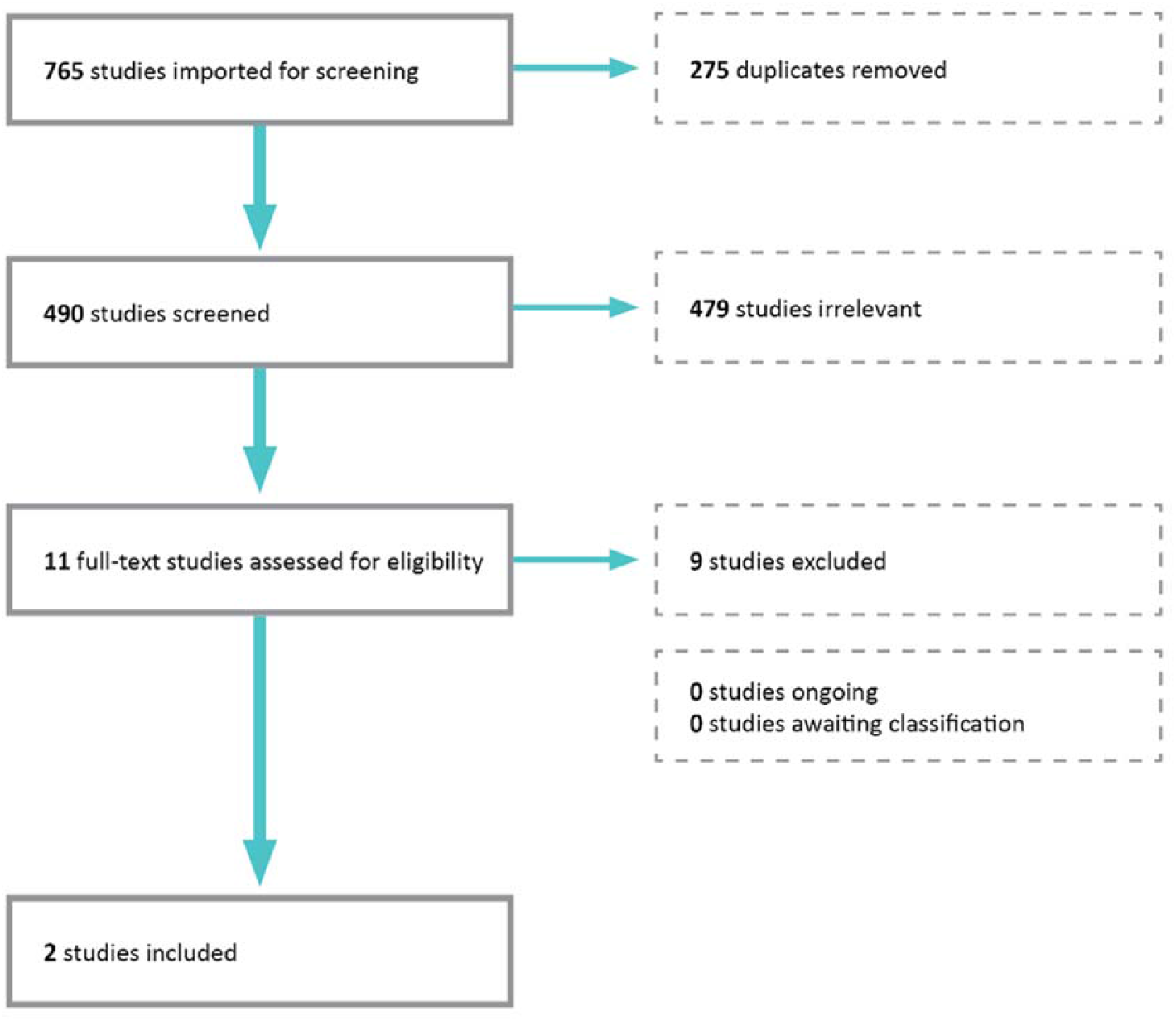
Prisma Flowchart

Both of the included studies were conducted in the UK, were prospectively registered and had Shirley Thomas as the lead author. The first study conducted by Thomas et al. published in 2012 reported on the Communication and Low Mood (CALM) trial^24^; the second study published by Thomas et al. in 2019 reported on the Behavioural Activation Therapy for Post-Stroke Depression (BEADS) feasibility trial^25^. The CALM trial recruited patients from hospital wards and stroke rehabilitation services^24^. The same approach was employed in the BEADS trial^25^, however this trial also used a community stroke database to identify participants. The CALM trial^24^ invited 129 participants with post-stroke aphasia, of which 105 agreed to participate. The BEADS trial^25^ invited 56 patients from three centres, of which 49 consented with the highest proportion of patients recruited through a community stroke database. The average number of BA sessions completed in the CALM trial^24^ was 9 out of a possible 20 sessions over a period of three months, with sessions lasting just under an hour on average (Table 1). In the BEADS trial^25^, the average number of BA sessions completed was 8.5 out of a possible 15 over a period of three months, and the average session length was also just under one hour (Table 1). Almost all the participants (92%) attended at least two BA sessions, with attendance slowly diminishing by the number of sessions – at least 5 sessions 88%, at least eight sessions 64% at least 10 sessions 40%.

**Table 1.** Overview of sessions for included studies.

| Name of trial | Average number of BA sessions | Average length of BA session |
| --- | --- | --- |
| BEADS<sup>25</sup> | 8.5 (range 0-14, SD 4.4) | 57 minutes, range 10–125 minutes, SD 13 minutes |
| CALM<sup>24</sup> | 9 (range 3–18, SD 2.63) | 58 minutes, range 30–89, SD 10.71 |

In both studies, assistant psychologists (APs are degree qualified roles within the National Health Service in the United Kingdom) were used to deliver BA. The BEADS trial^25^ reported the AP received training in BA through a two-day workshop. This included training from a speech and language therapist which is a modification of typical BA training to suit a post-stroke population. The CALM trial^24^ did not state whether the therapist received training in BA, however in a related paper^26^ that examined the treatment integrity of the intervention, it is noted that “training was also given in delivering behavioural therapy in accordance with the treatment manual” (p.1099). The length of the training was not reported. As was the case in the BEADS trial^25^, the CALM trial^24^ included training in supportive communication from a speech therapist. The APs in the CALM trial ^24^ attended joint monthly supervision and the therapy sessions were monitored through observation by the chief investigator. In the BEADS trial^25^, each therapist received weekly clinical supervision from a clinical psychologist, in addition to a monthly teleconference to discuss the interventions and any difficulties. Some therapy sessions were videorecorded and rated against the manual content to ensure fidelity^25^. As part of the BEADS trial^25^, APs were also given a manual that was revised and based on the CALM trial^24^.

Both studies used a manual that was a bespoke approach to BA for PSD. The CALM trial^24^ stated that the “manual had been developed from studies of cognitive behavioural therapy for depression after stroke and with older adults, and guidelines on conducting cognitive behavioural therapy with people with aphasia” (p. 400). The BEADS trial^25^ adapted the CALM manual to include “BA with stroke patients who do not have aphasia and provided examples and practical guidance relevant to all stroke patients” and incorporated “BA strategies used in low-intensity CBT and guidelines on adapting CBT for people with stroke” (p.38).

In the CALM trial^24^, 105 patients were randomized to either BA with usual care or TAU (usual care) alone; 51 patients were allocated to BA (48 participated in the BA intervention and 44 completed the BA program). In the BEADS trial^25^, 49 patients in total were randomized to either BA or TAU (described as “usual stroke care for treating people with post-stroke depression” p.vii); 26 patients were randomized to BA and 20 completed it. In the CALM trial^24^, there was no restriction on how long ago the patients had their stroke. The median post-stroke period for the TAU group was 9 months (interquartile range 4.9-39.0); the median post-stroke period for BA group was similar at 8.7 months (interquartile range 4.1-26.1). In the BEADS trial^25^, as part of the inclusion criteria, patients were a minimum of 3 months and a maximum of 5 years post stroke. Most of the participants in the BEADS trial experienced stroke 3 months to 1-year before the treatment (BA n=14, 64%; TAU n=14 60.9%). Other participants were either 1-2 years (BA n=7, 28%; TAU n=5, 21.7%) or 2-4 years poststroke (BA n=2, 8%; TAU n=4, 17.4%).

The CALM trial^24^ used the Stroke Aphasic Depression Questionnaire (SADQ v21) and the Visual Analog Mood Scale (VAMS); whilst the BEADS trial^25^ used the PHQ-9, with the SADQ as a secondary clinical outcome measure. The BEADS trial also used the VAMS. The CALM trial^24^ conducted post treatment assessments at 3 and 6 months, while in the BEADS trial^25^ only 6 months after baseline assessment was reported. Both studies noted improvements in the depression scores (that is, changes that indicate fewer depressive symptoms) in the BA group, and these were superior to those allocated to TAU. In the CALM trial^24^, Thomas et al. reported a decrease in depression scores (improvement in depression) from baseline to 3 months for both the BA group and the TAU group (Table 2). At the six month follow up, there was a slight increase from the 3 months scores. For the BA group, scores at six months remained below the baseline on the SADQ and the VAMS. However, for the TAU group, SADQ scores were above the baseline and VAMS scores were below the baseline. Regression analysis at three months suggested that group allocation alone was not a significant predictor for outcome measures, although when baseline values and communication impairment were controlled for, the effect of allocation became significant for the SADQ (P = 0.05) and VAMS (P = 0.03) (p.401). At six months, group allocation was a significant predictor of the SADQ (P = 0.045) and remained significant when baseline values were controlled for (P = 0.022). However, on the VAMS ‘sad’ item, there was no longer a significant effect of group allocation alone. In the BEADS trial^25^, Thomas et al. also reported a decrease in depression scores (that is, less depressive symptoms) from baseline to 6 month follow up for both the BA group and the TAU group, with decreases more notable in the BA group (Table 2). The authors noted small negative effects were found for the SADQ.

**Table 2.**
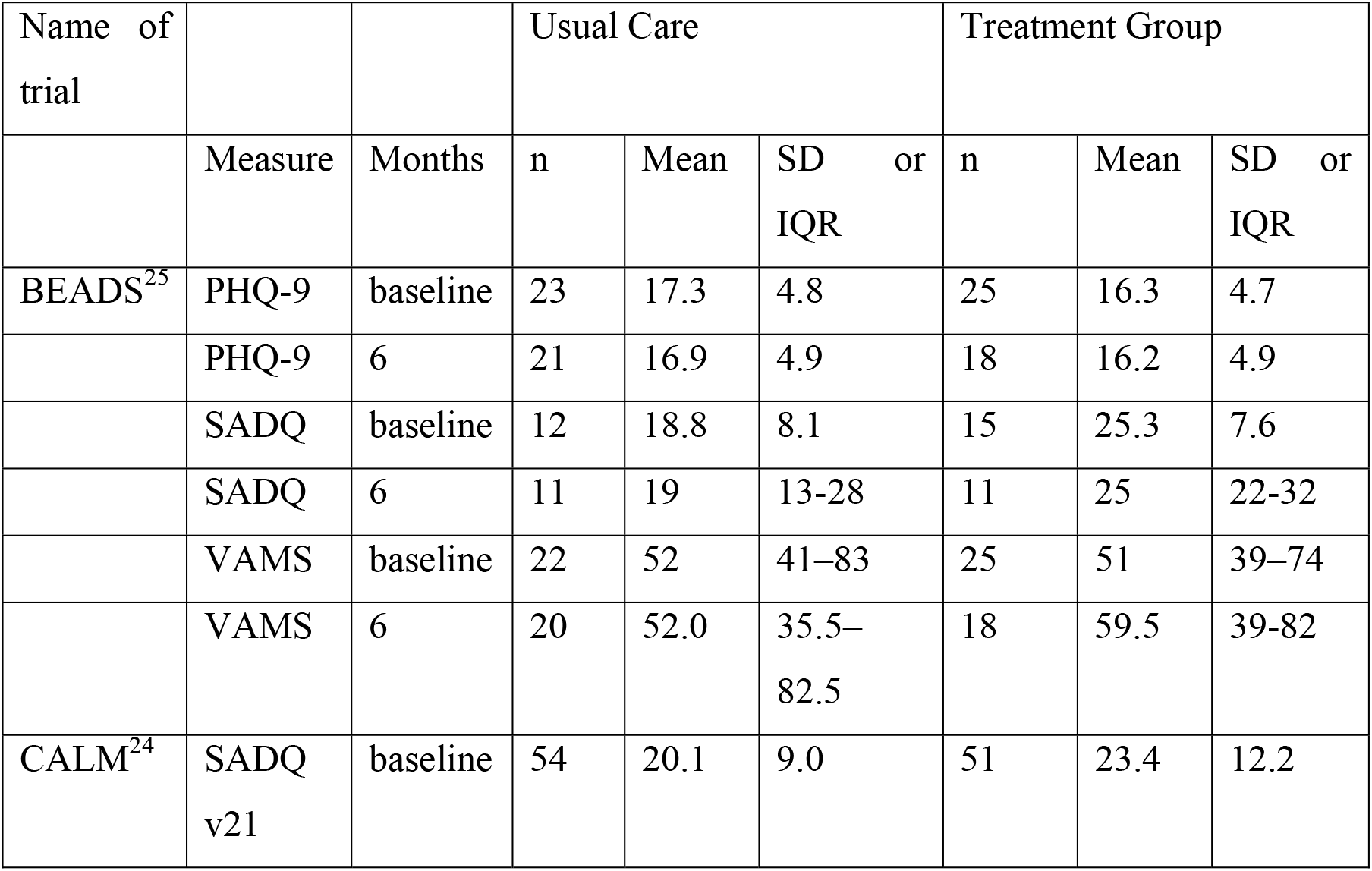
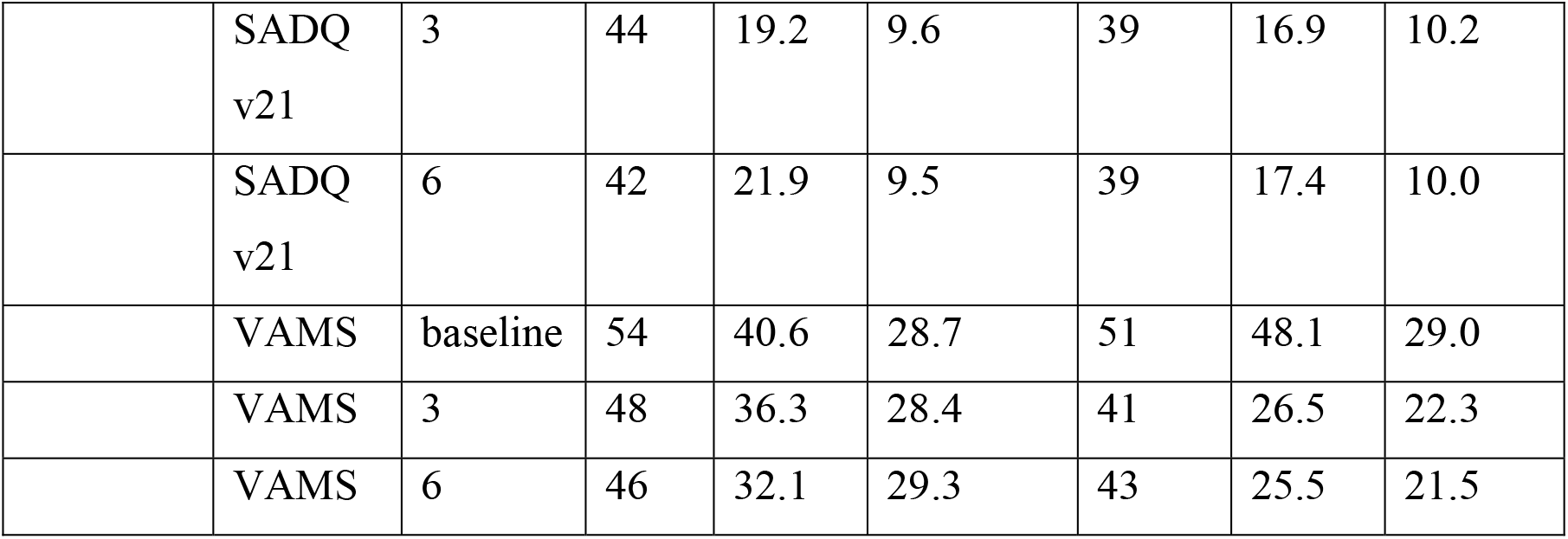
Patient outcomes by trial

Figure 2 shows the random effects meta-analysis results. Compared to treatment as usual, BA was associated with a decrease in depression symptoms in patients with post-stroke depression by a mean of 0.54 SD (SMD -0.54; 95%CI, -0.90 to -0.18).

**Figure 2.**
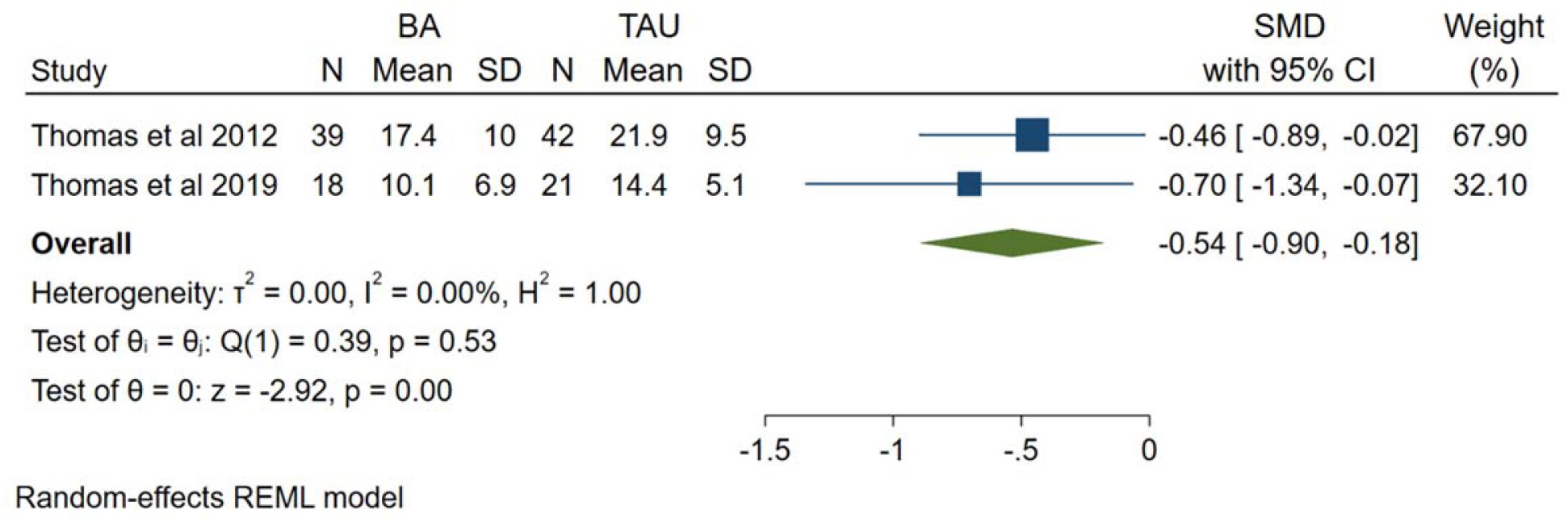
Meta-analysis showing the association between BA and PSD

We are unable to comment on the CALM trial with regard to harms or adverse events as there was no mention (that is, harms and adverse events were not reported in the paper)^24^. The BEADS trial^25^ reported on serious adverse events and adverse events. Three serious adverse events were reported, three individual participants were admitted to hospital or emergency following a suicide attempt, heart attack, and for a hernia repair. None of these events were considered related to the intervention. In terms of adverse events, 13 adverse events were experienced by 10 participants. Adverse events were suicidal intention, worsening health, fall, or a new health condition. Five of the adverse events were experienced by 4 participants in the intervention arm, while eight adverse events were experienced by six people in the control arm. Neither included suicide risk in the exclusion criteria.

### Risk of bias assessment

Of the two studies included in the review, one study was rated as having some concerns while the other study was rated as high risk of bias in the overall assessment of risk of bias (see Figure 3 and Figure 4).

**Figure 3.**
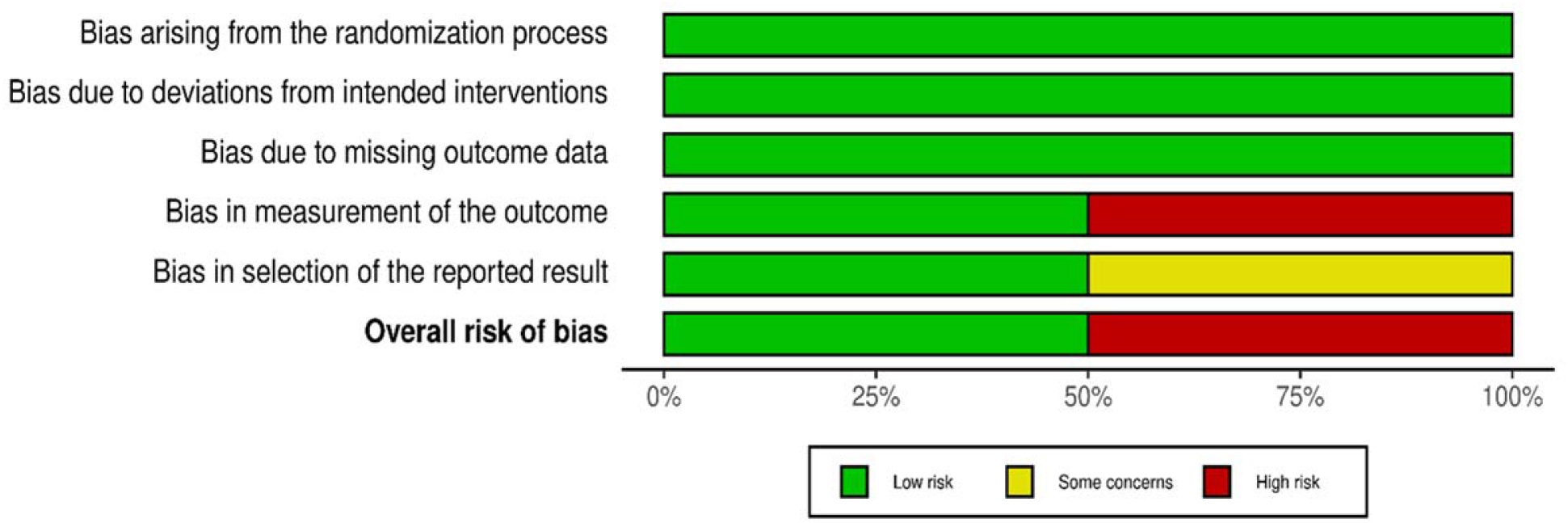
Risk of bias graph: review authors’ judgements about each risk of bias item presented as percentages across all included studies

**Figure 4.**
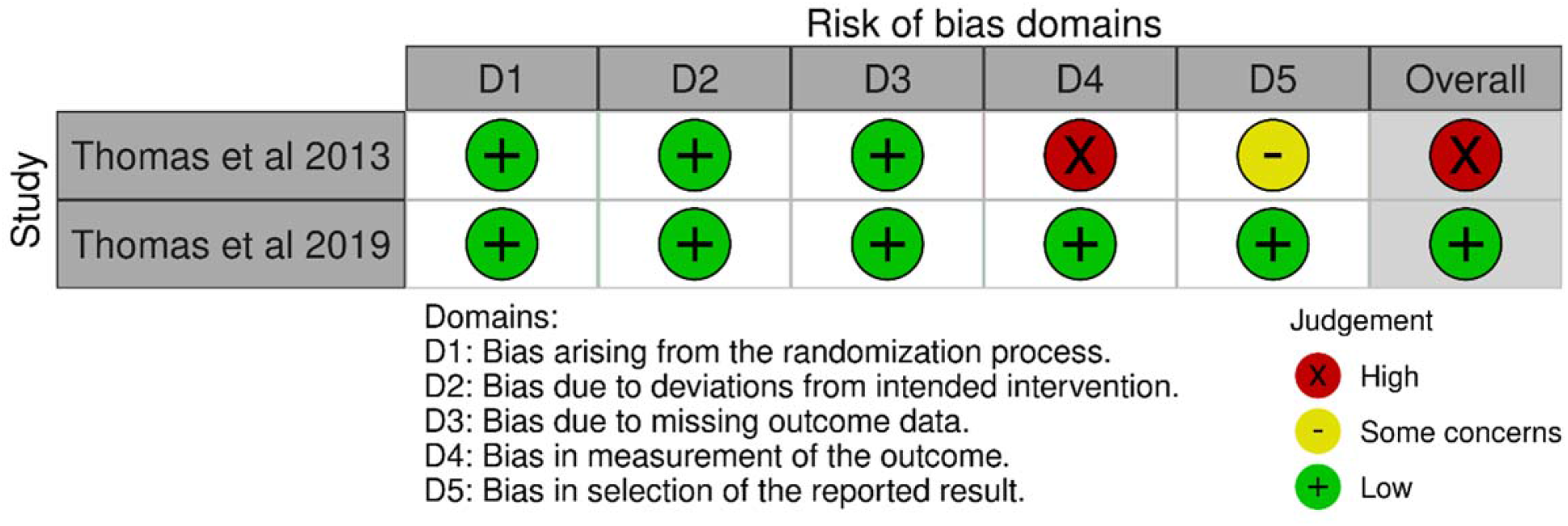
Risk of bias summary: review authors’ judgements about each risk of bias item for each included study

## Discussion

From these two studies there is some preliminary evidence that BA could be an effective intervention for reducing depressive symptoms in people with PSD. Our meta-analysis reported an SMD of 0.54, this is in comparison to Wang et al who reported a SMD of 0.76 in a meta-analysis of 23 RCTs examining the effectiveness of CBT in reducing PSD.

The results of the BEADS trial suggest decreases in scores on depressive scales from baseline to 6 months post intervention. While scores six months post intervention in the CALM trial remained improved compared to the baseline scores, there was a small increase from three months to six months. This is important as it may suggest that the effect of BA on PSD could potentially decline over time. This is something that future trials should consider in terms of the long term follow up of participants. Further, given that different challenges arise throughout the poststroke recovery journey, such as coping with grief and loss of function and independence, it may be important to consider whether another ‘dose’ of BA is required.

Completion and compliance to the BA program was promising and indicative that BA may be appropriate for poststroke patients. In the CALM trial, of the 48 people who participated in the BA program, 44 completed (91.67%). The completion rate in the BEADS trial was lower with 20 of the 26 participants randomized to the BA group completing the program (76.92%). The BEADS trial used compliance with BA as an outcome measure rather than as a covariate. They considered compliance to be whether participants attended the BA sessions. They note that attendance to therapy sessions was high. This preliminary evidence on that BA may be appropriate for the post-stroke population. Neither study reported the minimum therapeutic dose of BA required to treat PSD or whether more BA sessions resulted in improved outcomes. However, the potential complexity of PSD may indicate additional BA sessions are required.

As BA can be tailored to individual patients, it could also be suggested that BA would be acceptable to people with PSD. That is, a program of BA can be developed to accommodate physical needs associated with stroke. Notably, the CALM manual was adapted to include people with aphasia – this is an important point as people with communication difficulties post-stroke are often not considered in research evaluation. While the length of BA training was not mentioned in the CALM trial, the BEADS trial reported two-day BA training for therapists. In addition to BA training, both studies included training from a speech and language therapist that focused on communicating with stroke survivors who may have speech and/or language difficulties or cognitive impairment. It is unclear whether adaptions to the BA program itself were made to accommodate people with cognitive impairment. This is an important consideration as it could impact a stroke survivor’s ability to or motivation to engage in therapy. It may be necessary to take steps to overcome this, such as engaging with families and families to develop activities which aligns with the stroke survivor’s values, or conduct deeper analysis of the person’s history to identify previous activities which provided enjoyment which the person is still able to participate in.

In both studies, most patients were 3 months to 1 year post stroke. This is understandable as the most significant gains, physically and physiologically, are typically made in the six months post-stroke^27^. Equally, when the physical recovery from stroke becomes more prolonged, people may be faced with more psychological challenges, such as adapting to a new reality with different physical challenges that could obstruct activities they may have previously found rewarding. Adopting an early intervention approach for improving mood post stroke is an important consideration and is encouraged by clinical guidelines for post stroke depression^28^.

We note that in both studies an assistant psychologist delivered BA. Other studies have included different types of workers, including lay workers, to deliver BA for people with depression. An important consideration for people living in underserved areas, where people may be at higher risk of experiencing stroke and depression separately, is the access to mental health services. Training non-mental health professionals to deliver BA to people experiencing PSD – for example rehabilitation staff - may be a viable solution to this dilemma, increasing the reach of support. Emphasizing an early intervention approach could also reduce the need for access to specialist mental health professionals.

### Review limitations

In this review, we focused on a single primary outcome - depression. We could have included studies examining other outcome measures, such as quality of life, or the experiences of stroke survivors receiving BA in treating PSD. We restricted our review to English language journals and did not examine the grey literature. Hence, studies in this area may have been overlooked.

### Future research

The BEADS trial conducted by Thomas et al.^25^ established it is feasible to conduct a trial to test the association of BA and PSD. In both included studies, assistant psychologists delivered BA. Further research is required to test the feasibility and effectiveness of other groups of workers to deliver BA and what type of training and ongoing support these group of workers may require. This has particular relevance in countries where there is a maldistribution of health care professionals and services may be lacking in areas with greater need. Many PSD survivors are cared for in their community. It would be interesting to better understand how we can involve families in the delivery of BA. Such an approach may enhance the quality of a BA program for the PSD survivor and improve the health and wellbeing of the family.

## Conclusion

PSD can negatively impact stroke recovery and contributes to higher hospitalization, mortality, and reduced quality of life. Further, people with PSD are less likely to participate in rehabilitation programs that could improve their overall condition^4^. Given that approximately one third of people who experience a stroke will develop PSD, increasing access to psychological intervention may enhance clinical outcomes for people living with PSD. This systematic review and meta-analysis identified two peer-reviewed studies that examined BA with people with PSD. Both tested the effectiveness and feasibility of a BA delivered by an assistant psychologist and reported improvements in depression symptoms. As BA can be delivered effectively by non-specialist mental health workers, BA has the potential to reach larger numbers of people living with PSD, particularly those in underserved communities where there is increased risk of stroke.

## Data Availability

The data that support the findings of this study are available on request from the corresponding author.

## Acknowledgements

We thank Lorien Delaney, an Academic Librarian for her generous support and guidance.

## Author contribution

All authors conceived the study and were involved in the development of the protocol. ED developed the search strategy, drafted the protocol, registered it with OSF, and conducted database searches. ED, SW, and MJ performed title and abstract and full-text screening of retrieved papers. Any conflicts at full-text screening were resolved by RG. SW and MJ participated in data extraction. ED and MJ completed the quality appraisal of the included studies. EY performed the meta-analysis. SW and MJ prepared the draft versions of this manuscript. ED, SH, MG, and RG read the draft versions of the manuscript. All authors approved the present version of the manuscript. The data that support the findings of this study are available on request from the corresponding author (SW).

## Ethics statement

This review relied on previously published material and did not require ethical approval.

